# Associations of Health Insurance, Household Income, and Cost-Related Healthcare Barriers With Routine Medical Checkup Utilization Among U.S. Adults: Evidence From the 2024 BRFSS

**DOI:** 10.64898/2026.09.29.26364306

**Authors:** Chika Onwuameze, John Nwoha, Godyson Orji

## Abstract

**Objective:** To examine associations of health insurance status, household income, and cost-related healthcare barriers with receipt of a routine medical checkup among U.S. adults.

**Methods:** This cross-sectional study analyzed 2024 Behavioral Risk Factor Surveillance System data. Survey-weighted descriptive estimates and multivariable logistic regression incorporated sampling weights, strata, and primary sampling units. The outcome was a routine medical checkup within the previous 12 months.

**Results:** Among 321,592 complete cases, 78.0% of U.S. adults reported a recent routine checkup. Cost-related barriers (adjusted odds ratio [AOR], 0.56; 95% confidence interval [CI], 0.54-0.57) and uninsurance (AOR, 0.28; 95% CI, 0.27-0.29) were associated with lower checkup utilization. Lower household-income groups generally had lower adjusted odds than adults with incomes of $200,000 or more.

**Conclusions:** Insurance, income, and reported cost barriers were independently associated with routine checkup utilization. Findings are descriptive and do not establish causal effects.

## 1. Introduction

Routine medical checkups indicate health care utilization and provide opportunities for screening, counseling, vaccination, and risk assessment. Utilization, however, should not be treated as synonymous with access. Access is multidimensional and includes availability, accessibility, accommodation, affordability, and acceptability (Penchansky & Thomas, 1981). A routine checkup represents realized use of care rather than a measure of access. This distinction is important when examining financial factors that may shape routine care.

The relationship between insurance, financial exposure, and health care use is well established. Evidence from the RAND Health Insurance Experiment showed that greater cost sharing reduced health service use, while the Oregon Health Insurance Experiment demonstrated that gaining Medicaid coverage increased health care utilization and use of several preventive services (Baicker et al., 2013; Marino et al., 2016). More recent research continues to document financial barriers within insured populations. Hoagland et al. (2025) found unexpected cost sharing during preventive-care encounters, and Norris et al. (2022) reported that eliminating cost sharing generally increased preventive-service utilization, especially among financially vulnerable populations. These studies show that insurance coverage alone does not fully describe the financial conditions surrounding health care use.

Contemporary national surveillance remains useful because insurance status, household economic position, and reported cost-related barriers capture distinct financial circumstances. Recent U.S. studies have shown socioeconomic and racial or ethnic differences in wellness visits and preventive screening (Alba et al., 2024) and associations between accumulated social risks and lower routine-checkup use among low-income adults (Nguyen & Cole, 2025). In the Behavioral Risk Factor Surveillance System (BRFSS), the cost-related barrier item asks whether a respondent was unable to obtain needed medical care because of cost during the preceding 12 months. This measure is not specific to preventive care and should not be interpreted as a preventive-care access outcome. Here, it is treated instead as a self-reported indicator of experienced financial constraint and examined alongside insurance status and household income as an exposure associated with routine checkup utilization.

This study does not seek to establish a new causal effect of insurance or cost sharing. Rather, it provides national evidence on how these financial characteristics are associated with realized routine-care use in 2024. The objective of this study was to examine associations of health insurance status, household income, and cost-related healthcare barriers with receipt of a routine medical checkup within the previous 12 months among U.S. adults using 2024 BRFSS data.

## 2. New Contribution

This study provides contemporary national evidence on routine medical checkup utilization among U.S. adults using 2024 Behavioral Risk Factor Surveillance System data. Rather than treating insurance coverage, household income, and cost-related healthcare barriers as interchangeable measures of access, the analysis evaluates them as distinct financial characteristics associated with realized use of routine care. The cost-related barrier measure is used only as an indicator of reported financial constraint and is not interpreted as a preventive-care outcome. The study does not attempt to estimate causal effects or claim that the relationship between financial circumstances and healthcare use is new. Its contribution is to provide updated, population-level estimates of how insurance status, income, and reported cost barriers are independently associated with routine checkup utilization after adjustment for demographic and health characteristics. These findings offer current surveillance evidence on financial disparities in routine care use and identify population groups with lower utilization.

## 3. Methods

### 3.1. Study Design and Data Source

This cross-sectional study used the 2024 Behavioral Risk Factor Surveillance System (BRFSS), a telephone survey administered by the Centers for Disease Control and Prevention (CDC) in collaboration with state and territorial health departments. BRFSS collects information from noninstitutionalized adults aged 18 years or older on health status, health care use, insurance, chronic conditions, and health-related behaviors. The 2024 survey used probability-based landline and cellular telephone sampling. Analyses incorporated the complex survey design to generate population-representative estimates (Centers for Disease Control and Prevention [CDC], 2025a).

## 3.2. Study Population

The 2024 public-use file contained 457,670 respondents. Analyses were restricted to respondents with valid information for the outcome, primary exposures, and covariates included in the adjusted model. Responses coded as refused, “don’t know/not sure,” missing, or noninformative were treated as missing. The multivariable analysis used complete cases and included 321,592 respondents. Because the revised study focuses on one outcome—routine medical checkup utilization—the model previously treating cost-related inability to obtain needed care as a separate outcome was not retained.

### 3.3. Outcome

The sole outcome was receipt of a routine medical checkup within the previous 12 months. The BRFSS item assessing time since the most recent routine checkup was recoded as a binary variable indicating a checkup within the preceding year versus no checkup within that period. This measure was interpreted as health care utilization, not as a measure of health care access or receipt of all recommended preventive services. A routine checkup may create opportunities for prevention, but the item does not identify which preventive services were delivered.

### 3.4. Primary Exposures and Covariates

The exposures were health insurance status, household income, and a self-reported cost-related health care barrier. Insurance was categorized as insured or uninsured. Household income was grouped as less than $15,000; $15,000 to less than $25,000; $25,000 to less than $35,000; $35,000 to less than $50,000; $50,000 to less than $100,000; $100,000 to less than $200,000; and $200,000 or more. The highest-income group was selected as the reference category to permit consistent comparison of lower-income groups with the most economically advantaged category and to characterize the socioeconomic gradient in routine checkup utilization.

The cost-related barrier measure identified respondents who reported being unable to obtain needed medical care because of cost during the previous 12 months. Because this item concerns needed care generally rather than preventive care specifically, it was not treated as a preventive-care outcome. Instead, it was analyzed as a contemporaneous indicator of reported financial constraint. The overlapping 12-month recall periods for the cost-barrier and routine-checkup items preclude temporal ordering; associations were therefore interpreted as descriptive and noncausal.

Covariates were selected a priori based on established relationships with health care utilization and availability in BRFSS. The adjusted model included age, sex, race and ethnicity, educational attainment, employment status, marital status, body mass index, cigarette-smoking history, and diabetes status. Age and body mass index were modeled continuously.

### 3.5. Statistical Analysis

Survey-weighted descriptive statistics summarized the study population and estimated the prevalence of routine checkup utilization. A single survey-weighted multivariable logistic regression model estimated associations of insurance status, household income, and cost-related barriers with routine checkup utilization while adjusting for the prespecified covariates. The final respondent weight, stratification variable, and primary sampling unit were incorporated according to CDC analytic guidance (CDC, 2025b). Results are reported as adjusted odds ratios with 95% confidence intervals. Because routine checkup prevalence was high, odds ratios were interpreted strictly as odds ratios rather than risk or prevalence ratios. No causal interpretation was made. Analyses were conducted in R (R Foundation for Statistical Computing, Vienna, Austria).

### 3.6. Ethical Considerations

The study used publicly available, deidentified BRFSS data and involved no direct interaction with participants or access to identifiable private information. Institutional review board review was not required for this secondary analysis of public-use data.

## 4. Results

### 4.1. Sample Characteristics and Routine Checkup Utilization

The 2024 Behavioral Risk Factor Surveillance System included 457,670 adult respondents. The complete-case multivariable model of routine medical checkup utilization included 321,592 respondents. In descriptive analyses based on respondents with valid information for each characteristic, the survey-weighted mean age was 48.3 years (95% confidence interval [CI], 48.2-48.4), 50.9% were women, and 91.4% were insured. Non-Hispanic White adults represented 57.2% of respondents with valid race and ethnicity information, followed by Hispanic adults (19.6%), non-Hispanic Black adults (11.8%), other non-Hispanic racial groups (8.8%), and multiracial non-Hispanic adults (2.6%). The largest household-income category was $50,000 to less than $100,000 (28.4%) (Table 1).

**Table 1.** Selected Characteristics of U.S. Adults, 2024 Behavioral Risk Factor Surveillance System.

| Characteristic | Unweighted n | Survey-weighted estimate, % | 95% CI |
| --- | --- | --- | --- |
| Total respondents | 457,670 | — | — |
| Age, mean years | — | 48.3* | 48.2-48.4 |
| **Sex** |  |  |  |
| Male | 217,487 | 49.1 | 48.8-49.4 |
| Female | 240,183 | 50.9 | 50.6-51.2 |
| **Race and ethnicity** |  |  |  |
| Non-Hispanic White | 329,346 | 57.2 | 56.9-57.5 |
| Non-Hispanic Black | 35,172 | 11.8 | 11.6-12.0 |
| Other race, non-Hispanic | 24,917 | 8.8 | 8.5-9.0 |
| Multiracial, non-Hispanic | 10,486 | 2.6 | 2.5-2.7 |
| Hispanic | 48,646 | 19.6 | 19.3-19.9 |
| **Health insurance** |  |  |  |
| Insured | 413,720 | 91.4 | 91.2-91.6 |
| Uninsured | 25,406 | 8.6 | 8.4-8.8 |
| **Education** |  |  |  |
| Did not graduate high school | 27,149 | 11.2 | 10.9-11.4 |
| High school graduate | 115,877 | 27.4 | 27.1-27.7 |
| Some college or technical school | 120,732 | 29.7 | 29.3-30.0 |
| College or technical school graduate | 191,549 | 31.8 | 31.5-32.1 |
| **Household income** |  |  |  |
| Less than \$15,000 | 20,800 | 6.2 | 6.0-6.4 |
| \$15,000 to less than \$25,000 | 33,503 | 9.1 | 8.9-9.3 |
| \$25,000 to less than \$35,000 | 41,702 | 11.2 | 11.0-11.5 |
| \$35,000 to less than \$50,000 | 49,686 | 12.7 | 12.5-13.0 |
| \$50,000 to less than \$100,000 | 112,409 | 28.4 | 28.1-28.7 |
| \$100,000 to less than \$200,000 | 82,453 | 22.8 | 22.5-23.1 |
| \$200,000 or more | 29,694 | 9.6 | 9.3-9.8 |

Note. CI = confidence interval. Numbers are unweighted respondent counts; percentages, means, and 95% confidence intervals are survey weighted. Age is presented as the survey-weighted mean rather than a percentage. Estimates were calculated among respondents with valid information for each characteristic and therefore denominators vary across variables. Race and ethnicity estimates exclude respondents with missing or noninformative responses; the same approach was used for insurance, education, and household income. The estimates shown are descriptive characteristics of the 2024 BRFSS sample and should not be interpreted as characteristics of the 321,592 complete-case regression sample.

Overall, an estimated 78.0% of U.S. adults reported receiving a routine medical checkup within the previous 12 months (95% CI, 77.7%-78.3%). An estimated 12.3% reported being unable to obtain needed medical care because of cost during the same period. The latter measure was treated as an exposure indicating reported financial constraint rather than as an outcome of preventive-care access.

### 4.2. Financial Characteristics Associated With Routine Checkup Utilization

In the survey-weighted multivariable logistic regression model, adults reporting a cost-related health care barrier had lower adjusted odds of receiving a routine checkup than those who did not report such a barrier (adjusted odds ratio [AOR], 0.56; 95% CI, 0.54-0.57). Uninsured adults also had substantially lower adjusted odds of a routine checkup compared with insured adults (AOR, 0.28; 95% CI, 0.27-0.29) (Table 2 and Figure 1).

**Table 2.** Selected Adjusted Associations With Routine Medical Checkup Utilization Among U.S. Adults, 2024 BRFSS.

| Characteristic | AOR | 95% CI |
| --- | --- | --- |
| **Cost-related health care barrier** |  |  |
| Barrier versus no barrier | 0.56 | 0.54-0.57 |
| **Health insurance** |  |  |
| Uninsured versus insured | 0.28 | 0.27-0.29 |
| **Household income** |  |  |
| \$200,000 or more | 1.00 | Reference |
| \$100,000 to less than \$200,000 | 0.92 | 0.89-0.95 |
| \$50,000 to less than \$100,000 | 0.85 | 0.82-0.88 |
| \$35,000 to less than \$50,000 | 0.83 | 0.80-0.86 |
| \$25,000 to less than \$35,000 | 0.75 | 0.72-0.78 |
| \$15,000 to less than \$25,000 | 0.89 | 0.85-0.93 |
| Less than \$15,000 | 0.86 | 0.81-0.91 |
| **Selected demographic and health characteristics** |  |  |
| Female versus male | 1.60 | 1.57-1.63 |
| Non-Hispanic Black versus non-Hispanic White | 1.55 | 1.50-1.60 |
| Hispanic versus non-Hispanic White | 1.14 | 1.11-1.17 |
| Age, per additional year | 1.03 | 1.02-1.03 |
| Diabetes versus no diabetes | 3.28 | 3.13-3.43 |

Note. AOR = adjusted odds ratio; CI = confidence interval. The survey-weighted logistic regression model included health insurance, household income, cost-related health care barrier, race and ethnicity, age, sex, education, employment, marital status, body mass index, smoking history, and diabetes. The model included 321,592 complete cases. Estimates describe associations and should not be interpreted as causal effects. The \$200,000-or-more household-income group was used as the reference category to compare lower-income groups with the highest-income category.

**Figure 1.**
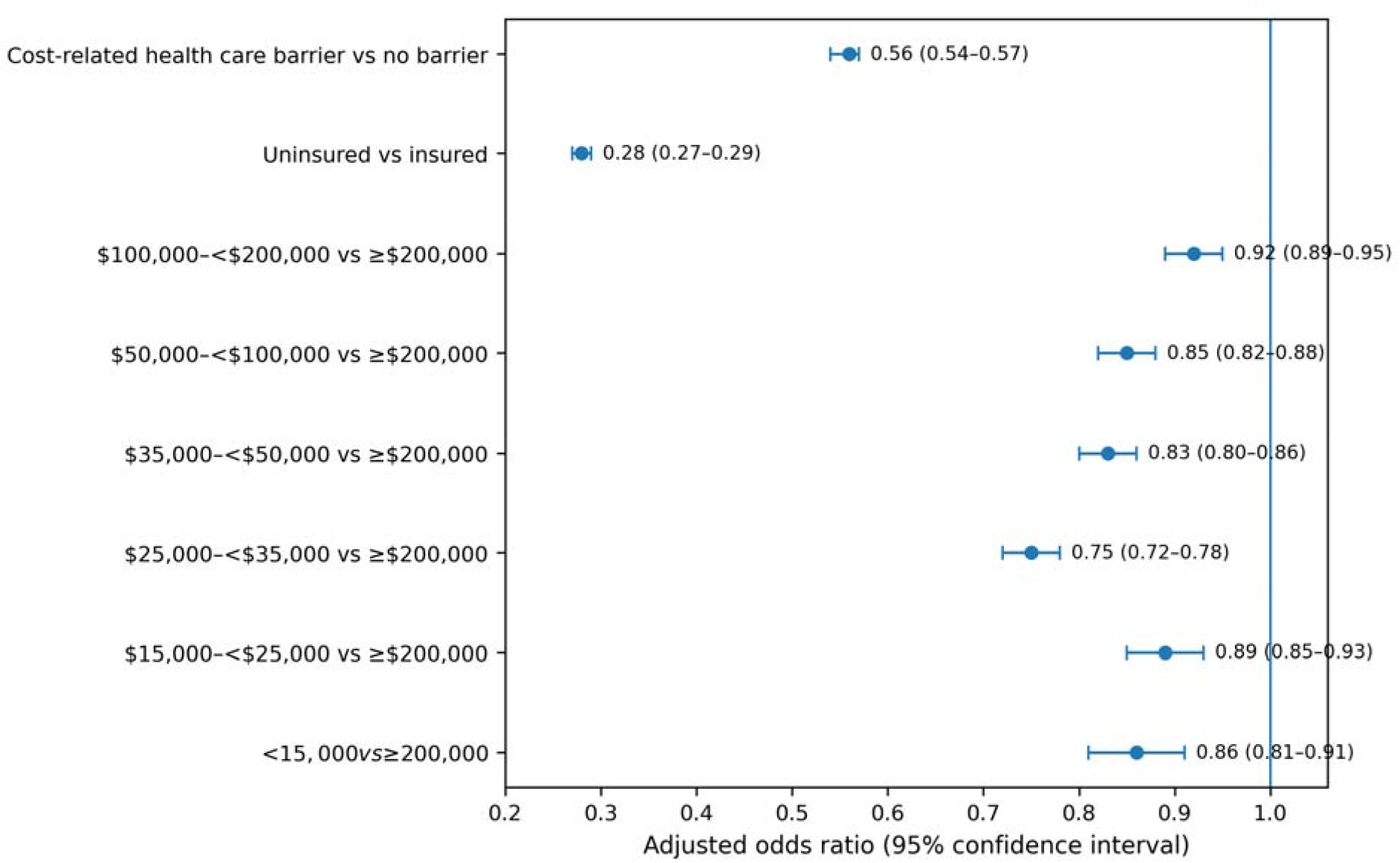
Adjusted associations of financial characteristics with routine medical checkup utilization among U.S. adults, Behavioral Risk Factor Surveillance System, 2024. Points represent adjusted odds ratios and horizontal bars represent 95% confidence intervals. The vertical reference line indicates an adjusted odds ratio of 1.00. The $200,000-or-more householdincome category is the reference group for income comparisons. Estimates are from a survey-weighted multivariable logistic regression model adjusted for health insurance, household income, cost-related health care barriers, race and ethnicity, age, sex, education, employment, marital status, body mass index, smoking history, and diabetes.

Household income was also associated with routine checkup utilization. Relative to adults with household incomes of $200,000 or more, adjusted odds were lower for those earning $100,000 to less than $200,000 (AOR, 0.92; 95% CI, 0.89-0.95), $50,000 to less than $100,000 (AOR, 0.85; 95% CI, 0.82-0.88), $35,000 to less than $50,000 (AOR, 0.83; 95% CI, 0.80-0.86), $25,000 to less than $35,000 (AOR, 0.75; 95% CI, 0.72-0.78), $15,000 to less than $25,000 (AOR, 0.89; 95% CI, 0.85-0.93), and less than $15,000 (AOR, 0.86; 95% CI, 0.81-0.91). The pattern was not strictly monotonic; the lowest adjusted odds occurred in the $25,000-to-lessthan-$35,000 category.

### 4.3. Demographic and Health Characteristics

Women had higher adjusted odds of receiving a routine checkup than men (AOR, 1.60; 95% CI, 1.57-1.63). Compared with non-Hispanic White adults, higher odds were observed among non-Hispanic Black adults (AOR, 1.55; 95% CI, 1.50-1.60) and Hispanic adults (AOR, 1.14; 95% CI, 1.11-1.17). Increasing age was associated with higher routine-checkup utilization (AOR per year, 1.03; 95% CI, 1.02-1.03), and adults with diabetes had higher adjusted odds than adults without diabetes (AOR, 3.28; 95% CI, 3.13-3.43).

All estimates represent cross-sectional associations and should not be interpreted as causal effects. Complete estimates from the revised single-outcome model are reported in the supplementary material.

## 5. Discussion

Using nationally representative 2024 Behavioral Risk Factor Surveillance System data, this study found that health insurance status, household income, and self-reported cost-related healthcare barriers were independently associated with routine medical checkup utilization among U.S. adults. Adults reporting a cost-related barrier had substantially lower adjusted odds of a routine checkup, and uninsured adults had markedly lower odds than insured adults. Lower household-income groups also generally had lower odds of routine checkup utilization than adults with household incomes of $200,000 or more, although the income pattern was not strictly monotonic. These findings should be interpreted as descriptive associations rather than causal effects.

The results are consistent with a substantial body of evidence showing that financial circumstances influence healthcare utilization. Experimental evidence from the RAND Health Insurance Experiment and the Oregon Health Insurance Experiment demonstrated that cost sharing and insurance coverage can affect healthcare use, including preventive services (Baicker et al., 2013; Marino et al., 2016). More recent research has shown that insured patients may still incur unexpected out-of-pocket costs for preventive care (Hoagland et al., 2025), while eliminating cost sharing generally increases preventive-service utilization, particularly among financially vulnerable populations (Norris et al., 2022). The present study does not establish a new causal relationship. Rather, it provides contemporary national estimates showing that insurance status, household income, and reported cost-related barriers remained independently associated with routine checkup utilization in 2024.

The cost-related barrier measure requires particularly careful interpretation. The BRFSS question asks whether cost prevented respondents from obtaining needed medical care during the previous 12 months. It does not identify whether the needed care was preventive, acute, chronic, mental health, or another type of service. Accordingly, the measure was used here only as an indicator of experienced financial constraint and not as an outcome or direct measure of preventive-care access. This distinction is important because access is multidimensional, whereas receipt of a routine medical checkup reflects healthcare utilization, or realized use of care (Penchansky & Thomas, 1981). Moreover, because the cost-barrier and routine-checkup questions both refer to the preceding 12 months, their temporal ordering cannot be determined.

Household income was also associated with routine checkup utilization after adjustment for insurance, cost-related barriers, and demographic and health characteristics. Nguyen and Cole (2025) similarly reported that accumulated social risks among low-income adults were associated with lower likelihood of routine checkups, while Alba et al. (2024) documented socioeconomic and racial or ethnic differences in wellness visits and preventive screening.

However, the income pattern in the present study was not strictly graded: the $25,000-to-less-than-$35,000 category had the lowest adjusted odds rather than the lowest-income category. This finding argues against interpreting household income as a simple dose-response measure and may reflect unmeasured differences in employment, insurance characteristics, health needs, healthcare preferences, or service availability.

Women, older adults, non-Hispanic Black adults, Hispanic adults, and adults with diabetes had higher adjusted odds of a recent routine checkup. These findings describe utilization of routine visits only and should not be interpreted as evidence that these groups have greater overall healthcare access or receive all recommended preventive services. A reported checkup indicates contact with healthcare but does not establish which preventive services were delivered, whether services were clinically indicated, or their quality.

This study has several strengths, including a large contemporary national sample and incorporation of survey weights, strata, and primary sampling units. The cross-sectional design precludes causal inference, and all principal measures were self-reported. Insurance was measured only as insured versus uninsured, preventing direct assessment of deductibles, copayments, benefit generosity, network adequacy, coverage continuity, or underinsurance.

Residual confounding by health literacy, healthcare preferences, provider availability, transportation, geographic accessibility, and other unmeasured factors is possible. Complete-case analysis may also introduce bias if missingness was systematic. Finally, because routine checkup prevalence was high, adjusted odds ratios should not be interpreted as risk or prevalence ratios.

These findings provide current national surveillance evidence that financial circumstances remain associated with routine medical checkup utilization. They support continued examination of insurance, household economic position, and reported cost-related barriers as distinct correlates of realized routine-care use while highlighting the need for longitudinal and quasi-experimental research to evaluate causal mechanisms.

## 6. Conclusion

This study found that health insurance status, household income, and self-reported cost-related healthcare barriers were independently associated with routine medical checkup utilization among U.S. adults in 2024. Uninsured adults and those reporting cost-related barriers had lower adjusted odds of a recent routine checkup, and lower-income groups generally showed lower utilization than adults in the highest-income category. These findings should be interpreted as descriptive associations rather than causal effects. The cost-related barrier measure reflects difficulty obtaining needed care because of cost and is not specific to preventive care; accordingly, it should not be viewed as a direct measure of preventive-care access. Instead, the results provide contemporary national evidence on financial correlates of realized routine-care use. Continued surveillance, together with longitudinal and quasi-experimental research, is needed to clarify mechanisms and determine whether changes in insurance design, affordability, or other financial conditions improve routine checkup utilization across population groups.

## Data Availability

The data used in this study are publicly available from the U.S. Centers for Disease Control and Prevention (CDC) through the 2024 Behavioral Risk Factor Surveillance System (BRFSS) public-use dataset. No restricted or proprietary data were used.

https://www.cdc.gov/brfss/annual_data/annual_2024.

## Data Availability

The 2024 Behavioral Risk Factor Surveillance System public-use data are publicly available from the Centers for Disease Control and Prevention.

