## Supplementary Table S1 for "Associations of Health Insurance, Household Income, and Cost-Related Healthcare Barriers With Routine Medical Checkup Utilization Among U.S. Adults: Evidence From the 2024 BRFSS"

**Supplementary Table S1. Survey-weighted multivariable logistic regression associations with routine medical checkup utilization among U.S. adults, Behavioral Risk Factor Surveillance System, 2024 (analytic sample = 321,592)**

Complete adjusted estimates from the single-outcome model reported in the main manuscript are shown below. The outcome was receipt of a routine medical checkup within the previous 12 months.

| **Variable** | **Adjusted odds ratio** | **95% confidence interval** |
| --- | --- | --- |
| Cost-related healthcare barrier: Yes | 0.56 | 0.54–0.57 |
| Insurance: Uninsured | 0.28 | 0.27–0.29 |
| Household income: $100,000 to <$200,000 | 0.92 | 0.89–0.95 |
| Household income: $50,000 to <$100,000 | 0.85 | 0.82–0.88 |
| Household income: $35,000 to <$50,000 | 0.83 | 0.80–0.86 |
| Household income: $25,000 to <$35,000 | 0.75 | 0.72–0.78 |
| Household income: $15,000 to <$25,000 | 0.89 | 0.85–0.93 |
| Household income: Less than $15,000 | 0.86 | 0.81–0.91 |
| Race/ethnicity: Black, non-Hispanic | 1.55 | 1.50–1.60 |
| Race/ethnicity: Other race, non-Hispanic | 0.99 | 0.96–1.03 |
| Race/ethnicity: Multiracial, non-Hispanic | 0.89 | 0.84–0.94 |
| Race/ethnicity: Hispanic | 1.14 | 1.11–1.17 |
| Age, per year | 1.03 | 1.02–1.03 |
| Sex: Female | 1.60 | 1.57–1.63 |
| Education: Some college/technical school | 0.89 | 0.87–0.91 |
| Education: High school graduate | 0.90 | 0.88–0.93 |
| Education: Did not graduate high school | 0.85 | 0.82–0.88 |
| Employment: Self-employed | 0.88 | 0.85–0.90 |
| Employment: Out of work ≥1 year | 0.83 | 0.78–0.87 |
| Employment: Out of work <1 year | 1.24 | 1.18–1.30 |
| Employment: Homemaker | 0.95 | 0.90–0.99 |
| Employment: Student | 1.64 | 1.57–1.72 |
| Employment: Retired | 1.79 | 1.72–1.87 |
| Employment: Unable to work | 2.14 | 2.03–2.26 |
| Marital status: Divorced | 0.85 | 0.82–0.87 |
| Marital status: Widowed | 0.88 | 0.84–0.93 |
| Marital status: Separated | 0.83 | 0.78–0.88 |
| Marital status: Never married | 0.88 | 0.85–0.90 |
| Marital status: Unmarried couple | 0.78 | 0.75–0.81 |
| Body mass index, per kg/m² | 1.02 | 1.02–1.02 |
| Smoking history: Yes | 0.88 | 0.86–0.89 |
| Diabetes: Yes | 3.28 | 3.13–3.43 |

*Note.* The model was survey weighted and accounted for the Behavioral Risk Factor Surveillance System complex sampling design. Adjusted odds ratios are reported with Wald 95% confidence intervals. Reference categories were no cost-related healthcare barrier; insured; household income of $200,000 or more; White, non-Hispanic race and ethnicity; male; college or technical school graduate; employed for wages; married; no smoking history; and no diabetes. Age and body mass index were modeled continuously. Because the study is cross-sectional, estimates are descriptive associations and should not be interpreted as causal effects. The cost-related healthcare barrier item refers to inability to obtain needed medical care because of cost and is not specific to preventive care.
